# BPA Improved the Cognitive Dysfunction of Patients with CTEPH

**DOI:** 10.64898/2026.05.06.26352610

**Authors:** Sugang Gong, Yuanyuan Sun, Jing He, Wenhui Wu, Qinhua Zhao, Panpan Liu, Jinling Li, Huiting Li, Cijun Luo, Hongling Qiu, Jian Xu, Jinming Liu, Lan Wang, Ping Yuan

## Abstract

**BACKGROUND:** Chronic thromboembolic pulmonary hypertension (CTEPH) is a severe and progressive condition characterized by dyspnea and fatigue. Our previous study reported cognitive impairment in pulmonary hypertension (PH) patients. However, balloon pulmonary angioplasty (BPA) capable of alleviating cognitive impairment in patients with CTEPH is largely unknown.

**METHODS:** This was a prospective study involving a total of 131 patients with CTEPH who underwent BPA at the Shanghai Pulmonary Hospital. We collected Mini-Mental State Examination (MMSE) and Montreal Cognitive Assessment (MoCA) questionnaires and examined plasma Aβ and phosphorylated-tau217 (p-tau217) levels to assess the cognitive function of patients with CTEPH between the pre-BPA and post-BPA stages.

**RESULTS:** Following BPA, patients exhibited improved cognitive performance, accompanied by reduced plasma levels of Aβ_1-42_ and p-tau217. After the third BPA session, patients with a mean pulmonary arterial pressure (mPAP) of≥25 mmHg had significantly lower MMSE and MoCA scores compared to those with an mPAP of <25 mmHg. Linear regression analyses revealed that baseline and post-intervention MMSE or MoCA total scores were significant predictors of cardiac output (CO) levels measured after the last BPA procedure. Logistic regression analyses incorporating pre- and post-BPA clinical parameters identified three independent predictors of baseline cognitive dysfunction: lower educational attainment, higher baseline Aβ_1-42_ levels, and elevated baseline p-tau217 concentrations.

**CONCLUSIONS:** Our findings suggest promising therapeutic effects of BPA, associated with improvements in cognitive dysfunction and reductions in plasma Aβ_1-42_ and p-tau217 levels in patients with CTEPH.

**NOVELTY AND RELEVANCE:** *What Is New?:* This is the first study to demonstrate that balloon pulmonary angioplasty (BPA) improves cognitive function (MMSE/MoCA scores) in patients with chronic thromboembolic pulmonary hypertension (CTEPH). And the first report that BPA reduces plasma levels of Aβ1-42 and p-tau217— key Alzheimer’s disease-related proteins—in CTEPH patients, establishing a peripheral biomarker for CTEPH-associated cognitive impairment.

*What Is Relevance?:* Cognitive impairment is common but underrecognized in CTEPH, BPA now addresses both cardiopulmonary and cognitive dysfunction, improving quality of life beyond hemodynamic recovery. Findings support the cardiopulmonary-brain axis in CTEPH: improved pulmonary hemodynamics and oxygenation reduce systemic pathological protein release, benefiting brain function.

*Clinical/Pathophysiological Implications?:* Our findings suggest promising therapeutic effects of BPA, associated with improvements in cognitive dysfunction and reductions in plasma Aβ_1-42_ and p-tau217 levels in patients with CTEPH.

---

Chronic thromboembolic pulmonary hypertension (CTEPH) is characterized by chronic thrombus formation in the pulmonary artery, causing elevated pulmonary arterial pressure and right heart overload, possibly leading to right heart failure and death in severe cases.^1,2^ Pathological specimens showed arteriopathy, microvascular thrombosis, intimal hyperplasia, and a decrease in pulmonary artery cross-sectional area caused by thrombosis. Therefore, symptoms may be mild initially but can worsen without treatment.^3^ Pulmonary endarterectomy (PEA) is an established treatment for these patients.^4^ Patients with inoperable CTEPH and patients with persistent or recurrent PH after PEA were treated with Riociguat or balloon pulmonary angioplasty (BPA).^1,5^ Recently, our group reported that the total scores of Mini-Mental State Examination (MMSE) and Montreal Cognitive Assessment (MoCA) tests were able to independently predict the risk of PH occurrence and the survival of patients with PH, implying cognitive impairment as a potential contributor in the etiology and progression of PH.^6^ Similar findings have been reported in an external study with a particular focus on PAH.^7^ Notably, the cognitive performance in these studies was evaluated by questionnaires, whether there is a cognitive improvement after BPA remains unraveled.

Cognitive impairment is associated with the deregulation of various pathogenic molecules in the brain and the periphery. Among these molecules, amyloid-beta (Aβ) and phosphorylated tau proteins (p-tau) proteins in the brain are the most common and key pathological factors of Alzheimer’s disease (AD) and related cognitive impairment.^8,9^ In AD, β-secretase and γ-secretase cleave amyloid precursor protein into Aβ peptides such as Aβ_1-40_ and Aβ_1-42_. Although Aβ_1-40_ is more abundant in the brain, Aβ_1-42_ is the major component of amyloid plaques that induce neurodegeneration and cognitive dysfunction.^10-12^ Tau is an axon-enriched microtubule-associated protein that is hyperphosphorylated and aggregates into paired helical filaments to form neurotoxic neurofibrillary tangles in the brain.^13,14^ With the help of the glymphatic system and the disruption of the blood-brain barrier (BBB) integrity under pathological conditions, Aβ and p-tau in the brain enter the bloodstream, leading to their presence in the plasma.^15-20^ Various studies have demonstrated plasma Aβ_1-42_ and phosphorylated-tau217 (p-tau217) as reliable and robust biomarkers for detecting and monitoring cognitive decline in individuals with AD and related conditions.^13,14,21-26^ However, few studies have been conducted to investigate the changes in cognitive function, Aβ_1-42,_ and p-tau217 protein levels in patients with CTEPH after BPA.

Here, we aimed to investigate the scores of the MMSE and MoCA, plasma Aβ1-42 levels, and p-tau217 levels in relation to CTEPH in the prospective study. We observed that after BPA, patients exhibited significantly higher MMSE and MoCA scores, along with lower plasma Aβ1-42 and p-tau217 levels; these changes were associated with the severity of CTEPH. Additionally, a follow-up of these patients revealed that scores on the MMSE and MoCA at baseline could predict cardiac output (CO) in CTEPH patients. Notably, educational, baseline plasma Aβ1-42 and p-tau217 levels could predict MMSE and/or MoCA scores at baseline. Our study highlights the significant differences in cognitive functions and plasma Aβ1-42 and p-tau217 levels between before and after BPA, suggesting that BPA may be a promising therapeutic approach for CTEPH-induced cognitive dysfunctions.

## METHODS

### Data Availability

All of the data, analytical methods, and corresponding materials used in this study are available from the respective authors upon reasonable request.

### Study Design and Population

This was a prospective study involving patients with a clinical diagnosis of CTEPH who received BPA from January 2021 to July 2025 in the Shanghai Pulmonary Hospital. We collected MMSE and MoCA questionnaires, as well as a blood sample, to assess the cognitive function of patients with CTEPH. A total of 131 patients with complete information were continuously enrolled. There were 42 patients lost to follow-up, censored as alive on the last day of contact. Clinical data of all patients were retrieved from electronic health records. This study was approved by the Ethics Committee of Shanghai Pulmonary Hospital (number: K23-206). Written informed consents were obtained from all participants.

### Treatment

Targeted CTEPH medications included ambrisentan, maxitantan, sildenafil, tadalafil, silepag, and riociguat. There were seven patients treated without the above drugs. There were 108 patients treated with riociguat monotherapy or with sildenafil/tadalafil or ambrisentan/maxitantan. There were 16 patients treated with medications other than riociguat.

### Ascertainment of Exposure – Measure of Aβ1-42 and p-tau217

We retrieved human plasma from patients during hospitalization and from the outpatient clinics for controls. The plasma levels of Aβ1-42 and p-tau217 were measured using ELISA kits according to the manufacturer’s instructions (Jianglai Biology, Shanghai, China).

### Ascertainment of Cognitive Function

The cognitive function was measured using MMSE^27^ and the MoCA tools^28^ at admission by trained nurses. MMSE is a widely used tool that evaluates multiple cognitive function domains, including orientation, memory, attention, language, and visuospatial skills. It consists of a series of questions and tasks to assess an individual’s cognitive status. The maximum score of MMSE is 30, and scores lower than 27 indicate cognitive function decline. The MoCA is another commonly used cognitive screening tool that assesses a broader range of cognitive domains, such as attention and concentration, executive functions, memory, language, visuospatial abilities, and orientation. MoCA has a maximum score of 30, with scores below 26 indicating potential cognitive impairment.

### Statistics

Scatterplots and Shapiro-Wilk tests were used to assess the normality of data distribution in age, 6-minute walk distance (6MWD), N-terminal pro-B-type natriuretic peptide (NT-proBNP), Aβ1-42, and p-tau217 levels. Normally distributed continuous variables were described as mean ± standard deviation (SD). For skewed continuous variables, median (interquartile range [IQR]) was used to describe distributions. Student t-tests were used to compare the differences in age and BMI between PH patients and controls with a normal distribution. The Mann-Whitney U test or Kruskal-Wallis analysis was used to compare differences between groups for nonparametric parameters, including plasma Aβ1-42 and p-tau217 levels. Descriptive statistics for categorical variables, including sex, classification, WHO functional class (WHO FC), and target therapy, were reported as frequencies/percentages, and their differences between groups were compared by Pearson χ2 or Fisher’s exact test.

A linear regression model was used to evaluate the sensitivity and specificity of the total scores of MMSE and MoCA, plasma Aβ1-42, and p-tau217 levels to predict the level of CO. We calculated the odds ratio (OR) and its 95% confidence interval (CI) by applying logistic regression models to assess the different cognitive functions. The correlations between MMSE and MoCA total scores and other clinical parameters were evaluated with Spearman’s rank correlations. A P value < 0.05 was considered statistically significant. Data analyses and figures were performed using SPSS 21.0, RStudio (version 3.5.0), and GraphPad Prism 9.

## RESULTS

### Baseline Characteristics

A total of 131 patients with CTEPH underwent BPA. Of these, 32.5% were male. Table 1 summarizes the demographic characteristics and hemodynamic parameters of the cohort. The mean (±SD) age for all CTEPH patients was 62.8 ± 11.1 years. Following BPA, significant hemodynamic and functional improvements were observed. Key parameters demonstrating significant increases included Body Surface Area (BSA), 6MWD, CO, Cardiac Index (CI), and Pulmonary Artery Oxygen Saturation (PA-SaO_2_). Conversely, significant decreases were noted in NT-proBNP, D-Dimer levels, WHO FC, mean Pulmonary Artery Pressure (mPAP), Pulmonary Vascular Resistance (PVR), echocardiography-derived Pulmonary Artery Systolic Pressure (PASP), Right Atrium (RA) area, and the Eccentricity Index (EI). Left Ventricular Ejection Fraction (LVEF) also showed a significant decrease. Collectively, these changes indicate a substantial improvement in cardiopulmonary function attributable to BPA intervention (Table 1).

**TABLE 1.** Baseline and After BPA Characteristics in Patients with CTEPH.

|  | Baseline<br>(n=131) | After BPA<br>(n=131) | *P* value |
| --- | --- | --- | --- |
| Age, yrs | 62.8 ± 11.1 | 62.8 ± 11.1 |  |
| Male, n(%) | 53(32.5) | 53(32.5) |  |
| BSA | 1.68±0.17 | 1.69±0.17 | .040 |
| 6MWD, m | 335.80±124.98 | 450.82±69.01 | <.001 |
| NT-proBNP, pg/mL | 1636.83±5781.01 | 153.49±233.27 | .001 |
| D_Dimer | 445.00±667.63 | 226.41±359.70 | <.001 |
| WHO FC, I/II/III/IV, n | 1/39/100/9 | 1/92/51/0 | .016 |
| Hemodynamics |  |  |  |
| mPAP, mm Hg | 43±11.36 | 25.56±6.12 | <.001 |
| mPAWP, mm Hg | 83.53±21.64 | 47.94±12.47 | .066 |
| PVR, Wood units | 8.84±4.34 | 3.23±0.90 | <.001 |
| CO | 4.59±1.32 | 5.57±1.21 | <.001 |
| CI, L/min/m<sup>2</sup> | 2.76±0.70 | 3.38±0.74 | <.001 |
| PA-SaO<sub>2</sub> | 61.48±7.66 | 86.89±65.85 | <.001 |
| Echocardiography |  |  |  |
| PASP, mmHg | 72.14±24.27 | 41.75±18.67 | <.001 |
| RA area, mm<sup>2</sup> | 20.25±7.348 | 14.80±4.90 | <.001 |
| LVEF, % | 75.19±8.38 | 73.63±8.62 | <.001 |
| EI | 1.25±0.26 | 1.024±0.063 | <.001 |

Data are presented as No. (%) or median (interquartile range) unless otherwise indicated. 6MWD = 6-minute walk distance; BSA = body surface area; CI = cardiac index; CO = cardiac output; EI = eccentricity index; LVEF = left ventricular ejection fractions; mPAP = mean pulmonary arterial pressure; mPAWP = mean pulmonary capillary wedge pressure; NT-proBNP = N-terminal pro-brain natriuretic peptide; PA-SaO<sub>2</sub> = pulmonary artery oxygen saturation; PASP = pulmonary artery systolic pressure; PVR = pulmonary vascular resistance; RA = right Atrium; WHO FC = World Health Organization Functional Class.

### The Changes in Cognitive Functions in CTEPH Patients after BPA

Cognitive function, as assessed by both the MMSE and the MoCA, demonstrated significant improvement in patients with CTEPH following BPA. Post-BPA, CTEPH patients exhibited a highly significant overall increase in MMSE scores (*P* < .0001; Fig 1A). This improvement was evident across most cognitive domains assessed by the MMSE, including orientation, attention and calculation (A and C), recall, repetition, 3-step (3S) command execution, reading, writing, and figure copying (all *P* < .05; Fig 1B). Similarly, MoCA scores showed a highly significant overall increase after BPA (*P* < .0001; Fig 1D). Significant enhancements were observed in multiple MoCA subdomains, encompassing visuospatial/executive function (VS total), naming, forward/backward digit span (F/B DigitSpan), visuospatial abilities (VA), serial 7s subtraction, repetition, verbal fluency, abstraction, and orientation (all *P* < .05; Fig 1E). Notably, combination therapy involving BPA with riociguat and other targeted pulmonary hypertension PH medications yielded further significant cognitive benefits. This regimen significantly increased MMSE scores compared to baseline/pre-treatment values (*P* < .001; Fig 1C). It also considerably increased MoCA scores (*P* < .01; Fig 1F). Collectively, these findings suggest that BPA, particularly when integrated with targeted medical therapy (including riociguat), is associated with substantial and broad improvements in cognitive function among CTEPH patients.

**Figure 1.**
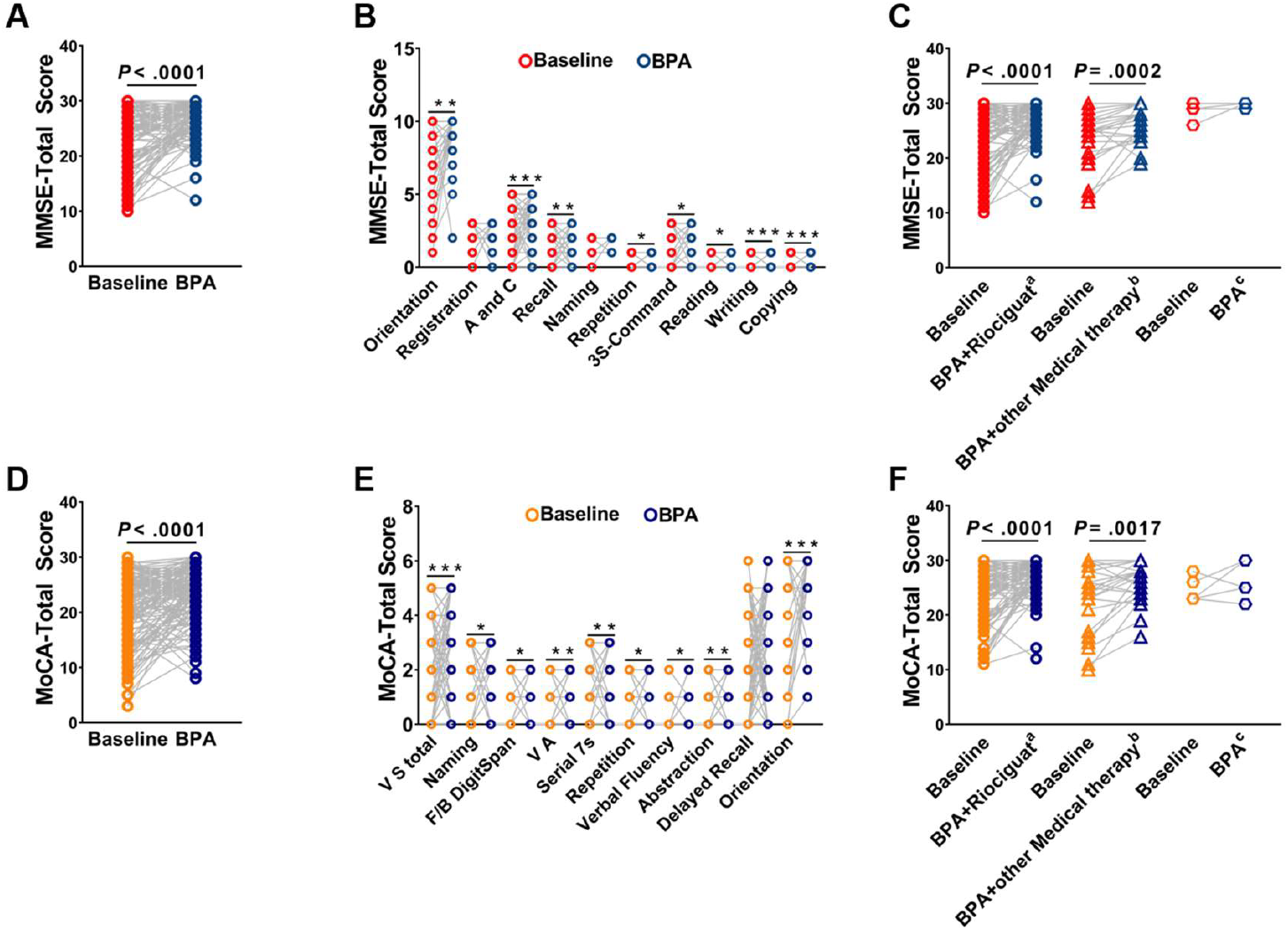
The changed of cognitive functions in CTEPH patients after BPA. **A**, MMSE-Total Score in pre-BPA patients of CTEPH. **B**, The cognitive domains assessed by the MMSE in pre-BPA and post-BPA. **C**, MMSE-Total Score among different treatment subgroups of CTEPH. **D**, MoCA-Total Score in pre-BPA patients of CTEPH. **E**, The cognitive domains assessed by the MoCA in pre-BPA and post-BPA. **F**, MoCA -Total Score among different treatment subgroups of CTEPH. A and C=attention and calculation, 3S=3-step, VS=visuospatial/executive function, F/B DigitSpan= forward/backward digit span, VA=visuospatial abilities. * < .005; ** < .001, *** < .0001.

### The Correlation between Cognitive Functions and Clinical Parameters in CTEPH Patients

At baseline, mild to moderate correlations—both positive and negative—were observed between patients’ MMSE or MoCA scores and key clinical parameters, including 6MWD, NT-proBNP levels, and CO (all *P* < .05; Fig 2A-F). Similarly, during follow-up assessments after the last BPA session, MMSE and MoCA scores demonstrated comparable mild to moderate positive or negative correlations with 6MWD, NT-proBNP, and CO (all *P* < .05; Fig 2G-L). Furthermore, linear regression analyses revealed that baseline MMSE and post-intervention MoCA and MMSE total scores served as significant predictors of CO measured after the last BPA procedure in CTEPH patients (Table 2).

**Table 2.** Linear Regression of MMSE and MoCA Total sScores for CO among CTEPH Patients with BPA.

| Overall association | OR | 95% CI | P value |
| --- | --- | --- | --- |
| **Last-BPA CO** |  |  |  |
| Baseline-MMSE total score | 0.052 | -0.027~0.051 | .555 |
| Baseline-MoCA total score | 0.196 | 0.005~0.071 | .024 |
| Last-MMSE total score | 0.196 | 0.005~0.154 | .037 |
| Last -MoCA total score | 0.209 | 0.007~0.101 | .026 |

BPA = balloon pulmonary angioplasty; CO = cardiac output; MMSE = mini-mental state examination; MoCA = montreal cognitive assessment

**Figure 2.**
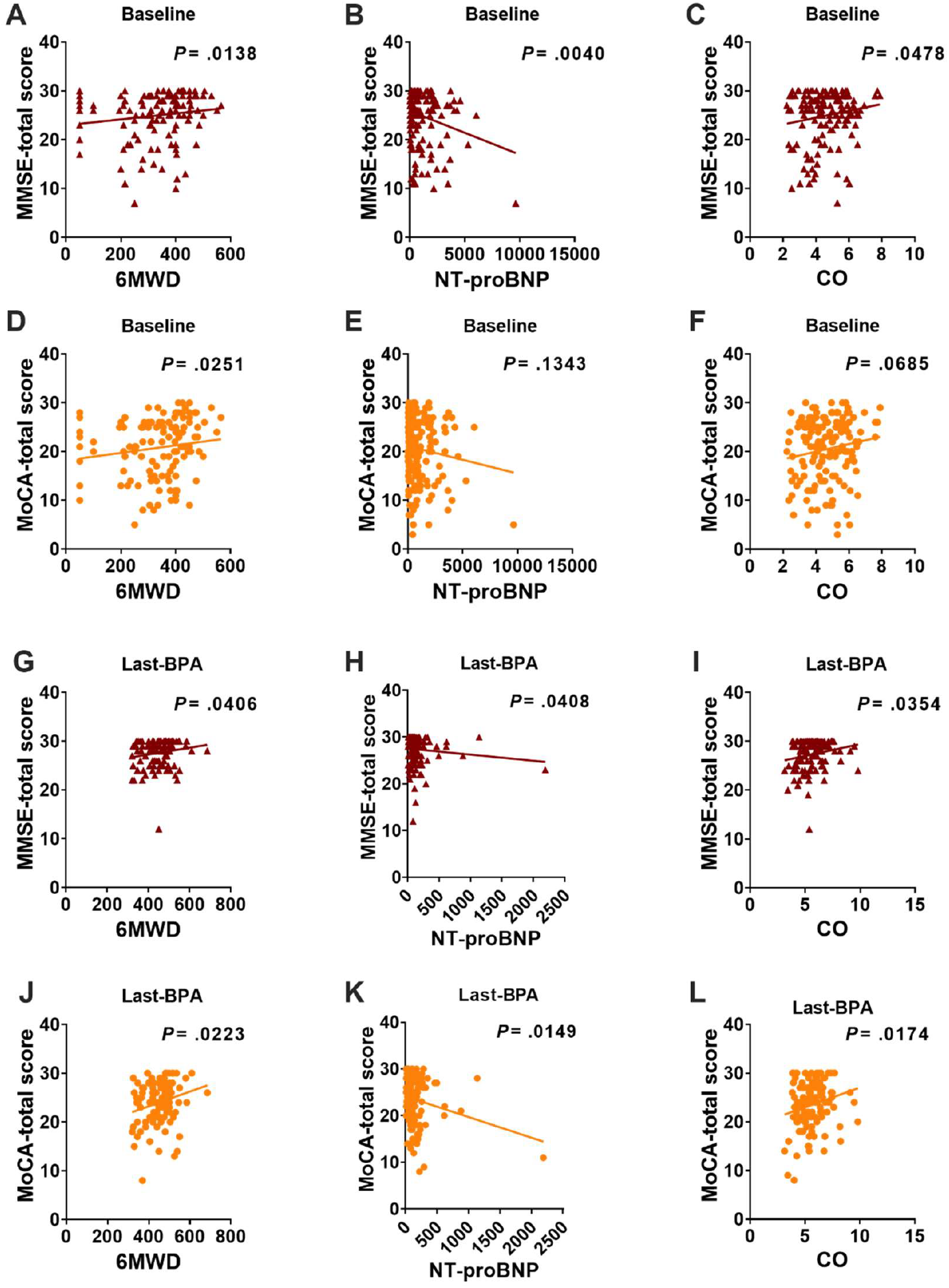
The correlation between cognitive functions and clinical parameters in CTEPH patients. **A-C**, The correlation between MMSE-Total Score and 6MWD, NT-proBNP, CO in baseline of CTEPH patients. **D-F**, The correlation between MoCA-Total Score and 6MWD, NT-proBNP, CO in baseline of CTEPH patients. **G-I**, The correlation between MMSE-Total Score and 6MWD, NT-proBNP, CO in the last BPA of CTEPH patients. **J-L**, The correlation between MoCA-Total Score and 6MWD, NT-proBNP, CO in last-BPA of CTEPH patients.

### Comparison of Cognitive Functions in CTEPH Patients after the 3rd BPA

Following the 3rd BPA session, patients exhibiting elevated mean pulmonary arterial pressure (mPAP ≥25 mmHg) demonstrated significantly lower MMSE and MoCA scores compared to those with lower mPAP levels (<25 mmHg) (both comparisons *P* < .05; Fig 3A-B). This inverse association between post-procedural hemodynamic burden and cognitive performance persisted despite the use of interventional therapy.

**Figure 3.**
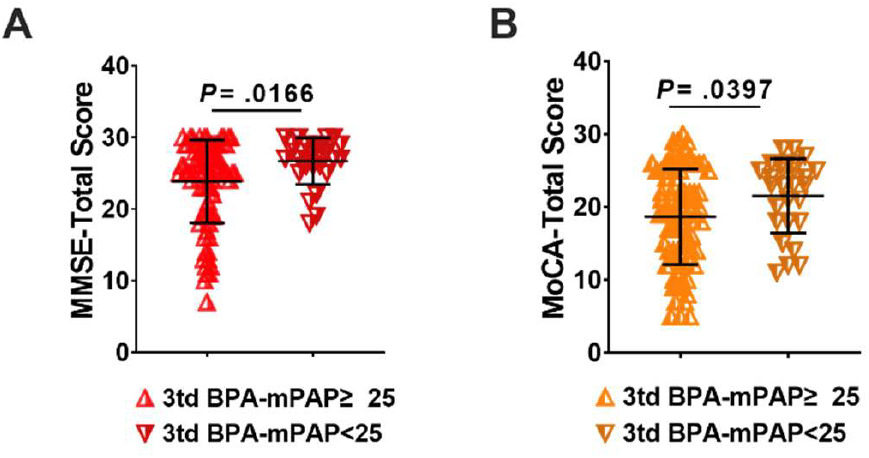
Comparison of cognitive functions in CTEPH patients after the third BPA. **A**, MMSE-Total Score in CTEPH patients with mPAP≥25 mmHg and <25 mmHg. **B**, MoCA-Total Score in CTEPH patients with mPAP≥25 mmHg and <25 mmHg.

### Comparison of Plasma Aβ1-42 and p-tau217 Levels in CTEPH Patients

Comparative analysis of plasma Aβ1-42 and p-tau217 levels revealed significant reductions in CTEPH patients after BPA (both *P* < .05; Fig 4A-B). Notably, elevated mPAP (≥25 mmHg) was consistently associated with higher Aβ1-42 and p-tau217 concentrations at both baseline and post-final BPA assessments compared to patients with mPAP <25 mmHg (*P* < .05, Fig 4C-D). While no significant intergroup differences in Aβ1-42 levels were observed across MMSE and MoCA score categories at baseline (Fig 4E), plasma p-tau217 levels were markedly elevated in patients with abnormal scores on both cognitive tests versus those with normal MMSE and MoCA scores, or with abnormal scores on only one test (both *P* < .05, Fig 4F).

**Figure 4.**
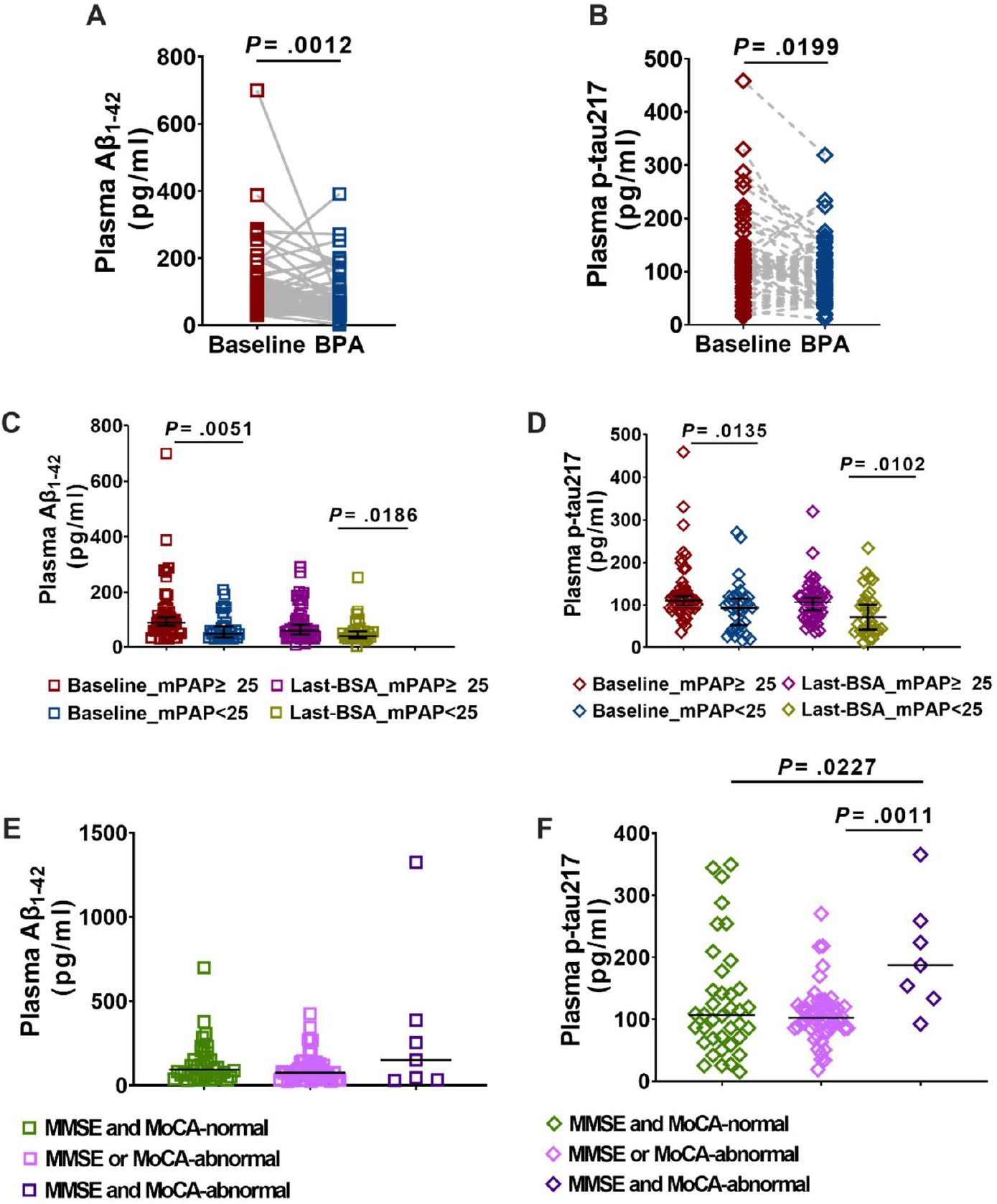
Comparison of plasma Aβ1-42 and p-tau217 levels in CTEPH patients. **A**, The plasma Aβ_1-42_ level in baseline and post-BPA of CTEPH patients. **B**, The plasma p-tau217 level in baseline and post-BPA of CTEPH patients. **C**, The plasma Aβ_1-42_ level in baseline and last-BPA of CTEPH patients with mPAP≥25 mmHg and <25 mmHg. **D**, The plasma p-tau217 level in baseline and last-BPA of CTEPH patients with mPAP≥25 mmHg and <25 mmHg. **E**, The Aβ_1-42_ levels in CTEPH patients with abnormal scores on both cognitive tests versus those with normal MMSE and MoCA scores, or with abnormal scores on only one test. **F**, The p-tau217 l levels in CTEPH patients with abnormal scores on both cognitive tests versus those with normal MMSE and MoCA scores, or with abnormal scores on only one test.

Multinational logistic regression analyses incorporating pre- and post-BPA clinical parameters identified three independent predictors of baseline cognitive dysfunction (defined as MMSE and/or MoCA abnormality): lower educational attainment, higher baseline Aβ1-42 levels, and elevated baseline p-tau217 concentrations (All *P* < .05). However, these factors did not predict cognitive status following the final BPA procedure. And for post-BPA patients, lower educational attainment, shorter post-6MWD, and lower CO were identified as independent predictors of cognitive dysfunction (Table 3).

**Table 3.** Multinational Logistic Regression of Parameters for Cognitive Function among CTEPH Patients with Before and After BPA.

| Overall association |  | OR | 95% CI | P value |
| --- | --- | --- | --- | --- |
| **Reference category--MMSE and MoCA-abnormal** |  |  |  |  |
| **MMSE and MoCA-normal** | Educational level | 0.005 | 0.000~0.056 | .005 |
| | Baseline Aβ<sub>1-42</sub> level | 0.997 | 0.994~1.001 | .108 |
|  | Baseline p-tau<sub>217</sub> level | 0.991 | 0.982~1.000 | .041 |
| **MMSE or MoCA-abnormal** | Educational level | 0.005 | 0.000~0.056 | .005 |
| | Baseline Aβ<sub>1-42</sub> level | 0.994 | 0.990~0.999 | .023 |
|  | Baseline p-tau<sub>217</sub> level | 0.986 | 0.977~0.995 | .003 |
| **MMSE and MoCA-normal** | Educational level | 0.005 | 0.000~0.056 | .005 |
|  | Last-BPA CO | 2.132 | 1.078~4.217 | .030 |
|  | Last-BPA 6MWD | 1.014 | 1.001~1.028 | .036 |
| **MMSE or MoCA-abnormal** | Educational level | 0.238 | 0.031~1.086 | .238 |
|  | Last-BPA CO | 2.009 | 1.022~3.950 | .043 |
|  | Last-BPA 6MWD | 1.010 | 0.997 ~1.023 | .141 |

MMSE = mini-mental state examination; MoCA = montreal cognitive assessment

## DISCUSSION

We analyzed 131 patients with CTEPH who underwent BPA, and comprehensively evaluated the impact of BPA on the cognitive impairment of patients with CTEPH, as well as its potential plasma biomarkers (Aβ1-42 and p-tau217) for the first time. Our results showed that BPA could significantly improve the cognitive impairment of CTEPH patients. After receiving BPA, the total scores of the MMSE and MoCA in patients increased compared to before BPA, and several parameters, including 6MWD, NT-proBNP, mPAP, PVR, and CO, also improved. This finding extends the therapeutic effect of CTEPH from the traditional cardiopulmonary hemodynamic to the cognitive domain, which is of great significance for disease diagnosis and treatment. A previous study has shown that among 25 CTEPH patients who underwent pulmonary endarterectomy (PEA) surgery, neurological complications occurred in 3 patients during the perioperative period. However, there was no significant difference in MMSE and MoCA scores before and after surgery,^29^ which differs from the results of our study. This may be related to the fact that our follow-up period for cognitive function was long, and the amount of data collected was substantial. In the future, further verification is needed through multi-center studies with a larger sample size. PH is associated with cognitive impairment, and this is attributed to chronic cerebral hypoperfusion, hypoxemia, and neurohumoral activation, etc. Our results further indicate that by relieving the obstruction of the pulmonary artery through BPA, reducing PVR and mPAP, and improving CO and systemic oxygen delivery (such as an increase in PA-SaO_2_), the microenvironment of the brain may be improved, allowing the patient’s cognitive impairment to recover. It is particularly noteworthy that the combined use of targeted drugs such as riociguat further enhances the cognitive benefits of BPA, suggesting that the effect of drug treatment in improving endothelial function and reducing vascular remodeling may provide additional protection for the brain.

Another significant contribution of our study is that, for the first time, we reported that BPA can reduce the levels of core proteins (Aβ1-42 and p-tau217) related to AD in the plasma of patients with CTEPH. This discovery has opened a new perspective for understanding the mechanism of PH-related cognitive impairment. Traditionally, Aβ and p-tau have been regarded as specific pathological products of central nervous system diseases, such as AD. However, Aβ levels increase in the cerebrospinal fluid of AD patients, their expression in plasma is decreased.^30-32^ However, recent discoveries have identified an important role for tau that is expressed in lung capillary endothelia. This endothelial tau stabilizes microtubules necessary for barrier integrity, yet infection drives production of cytotoxic tau and Aβ that are released into the circulation, where they contribute to end-organ dysfunction.^33^ In CTEPH patients, we found that the levels of Aβ1-42 and p-tau217 in plasma were elevated, which may reflect the pathological activities of Aβ and p-tau in the pulmonary vascular endothelial cells in CTEPH patients by chronic inflammation, hypoperfusion, and hypoxia, as well as the dysfunction of the endothelial barrier integrity. After BPA treatment, with the improvement of cardiopulmonary function, hemodynamics and oxygenation were optimized, which may promote the stability of the endothelial function, thereby leading to a decrease in the levels of these pathological proteins in the peripheral blood. This is consistent with the observed trend of cognitive function improvement, providing molecular-level evidence for the interaction between the “cardio-pulmonary-brain axis”. Of course, more data and deeper mechanism exploration are needed to prove this result further.

In addition, we found that for patients with mPAP ≥ 25 mmHg, after 3rd BPA treatments, their MMSE and MoCA total scores improved more significantly compared to patients with mPAP < 25 mmHg. This result strengthens the close connection between hemodynamic status and cognitive function. And the results of linear regression analysis suggest that baseline and postoperative MMSE/MoCA scores are significant predictors of CO levels after the last BPA treatment, indicating that cognitive function assessment may serve as a simple, non-invasive indicator that indirectly reflects the cardiac output status and overall hemodynamic recovery of patients, and has potential practical value in clinical follow-up. Through logistic regression analysis, we identified three independent predictors of baseline cognitive dysfunction in CTEPH patients: lower education level, higher baseline plasma Aβ1-42, and p-tau217. This further indicates the potential of Aβ1-42 and p-tau217 as biomarkers for cognitive impairment related to CTEPH.

Our study also has limitations, of course. Firstly, this is a single-center retrospective study. Although the sample size is acceptable, future studies should conduct larger samples, multi-center, prospective, and long-term research to verify the results of this study. Secondly, we measured the levels of Aβ and p-tau in plasma rather than in cerebrospinal fluid. Although plasma biomarkers have a more promising clinical translation potential due to their non-invasive nature, and numerous studies have confirmed their correlation with AD, their direct association with central nervous system pathology is still not as strong as cerebrospinal fluid testing. Future research, combined with neuroimaging (such as PET or MRI), will be able to reveal changes in brain structure and function more directly. Finally, this study did not explore the specific mechanism by which BPA improves cognitive function. Deeper mechanism exploration in the future may provide a better explanation for the results of this study.

## PERSPECTIVES

In summary, plasma Aβ1-42 and p-tau217 show promise as biomarkers for evaluating cognitive impairment related to CTEPH and monitoring treatment response. More importantly, our study provides robust evidence that BPA is an effective therapeutic strategy capable of improving cardiopulmonary function and cognitive performance. This enhanced understanding of CTEPH as a systemic disease establishes the central role of BPA in its treatment and offers new insights and hope for improving the quality of life and long-term prognosis of patients.

## Author Contributions

P.Yuan had full access to all the data in the study and take responsibility for the accuracy of the data analysis. S.Gong, L.Wang, and P.Yuan had the idea for and designed the study. J.Liu and P.Yuan supervised the study. Y.Sun did the statistical analysis, wrote the draft manuscript, and revised the manuscript. P.Yuan, S.Gong, Y.Sun, J.He, W.Wu, Q.Zhao, P.Liu, J.Li, H.Li, C.Luo, H.Qiu, J.Xu, J.Liu, and L.Wang contributed to the acquisition, analysis, or interpretation of data, and revised the manuscript. P.Yuan affirm that the manuscript is an honest, accurate, and transparent account of the study being reported. All authors revised the manuscript and approved the final version before submission.

